# Situation of COVID-19 in Brazil: An analysis via growth models as implemented in the ModInterv system for monitoring the pandemic

**DOI:** 10.1101/2021.03.29.21254542

**Authors:** Giovani L. Vasconcelos, Gerson C. Duarte-Filho, Arthur A. Brum, Raydonal Ospina, Francisco A. G. Almeida, Antônio M. S. Macêdo

## Abstract

In this work we analyze the cumulative curves of deaths attributed to COVID-19 in the 26 Brazilian States and the Federal District up until August 21, 2020. Mathematical growth models implemented by the application `ModInterv COVID-19`, which can be accessed via internet browser or via a mobile app, were used to investigate at which stage the epidemic is in each of the Brazilian federal units. The analysis revealed that almost all states in the Northern and Northeastern regions were already in the saturation phase, meaning that the epidemic was relatively under control, whereas in all Southern states and in most states in the Midwest the epidemic was still accelerating or showed only a slight deceleration. The Southeastern region presented a great diversity of epidemic stages, with each state being found at a different stage, ranging from acceleration to saturation. It is argued that understanding this heterogeneous geographical distribution of the epidemic is relevant for public health authorities, as it may help in devising more effective strategies against the COVID-19 pandemic in a continental country like Brazil.

## 1 Introduction

In Brazil, after more than five months since the first case of infection by the new coronavirus (SARS-CoV-2) and the first death from the disease (COVID-19) caused by the virus, the cumulative numbers of cases and deaths attributed to the disease showed no sign of stagnation. At the time of the writing of the first version of this paper, namely, in late August 2020, Brazil had more than 3.5 million confirmed cases and more than 110,000 deaths due to the disease [9, 13]. Looking at the situation of the epidemic, state by state, it was possible to observe that some of them seemed to be in a phase where the cumulative numbers of cases as well as of deaths still presented an accelerated growth, while other states seemed to have reached a stage where the numbers were growing more slowly or were already in a stage of saturation. In other words, there was a strong geographical heterogeneity of the epidemic in Brazil. In this complex context, it is therefore important to know the evolution of the COVID-19 epidemic in each of the federal units, since local governors had the legal autonomy to decide on the measures to be adopted to counter the propagation of the virus within their jurisdictions. Understanding the differences and similarities of the epidemic trajectories in the several regions of a continental country like Brazil may also help government and public health authorities to devise more effective strategies against the COVID-19 pandemic.

In this paper, we present an overview of the evolution of the COVID-19 epidemic in the 26 states of Brazil and the Federal District, up to August 21, 2020. In our analysis, we employed mathematical growth models to fit the cumulative curves of deaths attributed to COVID-19 in each federal unit. From the predictions of the model that best fits the data for a given geographical unit, it is possible to determine the evolution stage of the epidemic within the respective federal unit.

The models discussed in this work are implemented in the application `ModInterv COVID-19` [1], developed by our Research Network on Modeling the COVID-19 Epidemic and Non-Pharmacological Interventions–MODINTERV-COVID19 (http://fisica.ufpr.br/redecovid19). The application is available for general use via the internet (http://fisica.ufpr.br/modinterv) or via an Android mobile app available at the `Play Store` (https://play.google.com/store/apps/details?id=com.tanxe.COVID_19). In the `ModInterv` application, it is possible to monitor the cumulative and daily curves of cases and deaths of COVID-19 for all countries worldwide as well as for the states and cities in Brazil and the United States. In the present work we use the `ModInterv` to obtain an overview of the COVID-19 situation in Brazil up to mid-August, 2020, when all five stages of the COVID-19 epidemic defined in the `ModInterv` were still represented in Brazil. In other words, this date corresponds to a period of maximum diversity in the evolution patterns of the COVID-19 epidemic in Brazil. Hence it is an important epidemiological moment that deserves to be described and understood.

## 2 Data

In the present work, we analyze the cumulative curves of deaths attributed to COVID-19 for the 26 states of Brazil and the Federal District, with data updated until the date of August 21, 2020. The data used in our analysis are automatically “downloaded” from the website COVID19br.wcota.me, which in turn compiles and updates the data from the bulletins published by the respective State Health Departments [3].

The mortality data are normally tabulated according to the day on which each death occurred. Thus, an epidemic curve for a given State represents the accumulated number of deaths, *C*(*t*), attributed to COVID-19 that occurred in that State up to the time *t*, counted in days since the first death. It is inevitable that there may be some level of uncertainty in the data collection, such as under-reporting or delay in confirming deaths, but in general the data on deaths are considered more reliable than the data for infection cases, which are admittedly much more affected by the under-reporting problem, since many infections go undetected. For this reason, we have decided to restrict our analysis to the COVID-19 death curves.

As already mentioned, here we consider the fatality curves of each Brazilian federal unit up to August, 21, 2020. This upper limit date was chosen for our study because it corresponds to a period in which all five stages of the COVID-19 epidemic were still represented in Brazil; see below.

## 3 Methodology

### 3.1 Epidemic Curves and Stages

A typical epidemic curve [4], representing the cumulative number, *C*(*t*), of deaths as a function of the time *t*, counted in days from the day of the first death, is illustrated in Fig. 1(a). As indicated in this figure, the epidemic curve has two main phases, namely: i) a phase of accelerated growth, that is, with positive acceleration; and ii) a phase with decelerated growth, where the acceleration is negative.

**Fig. 1:**
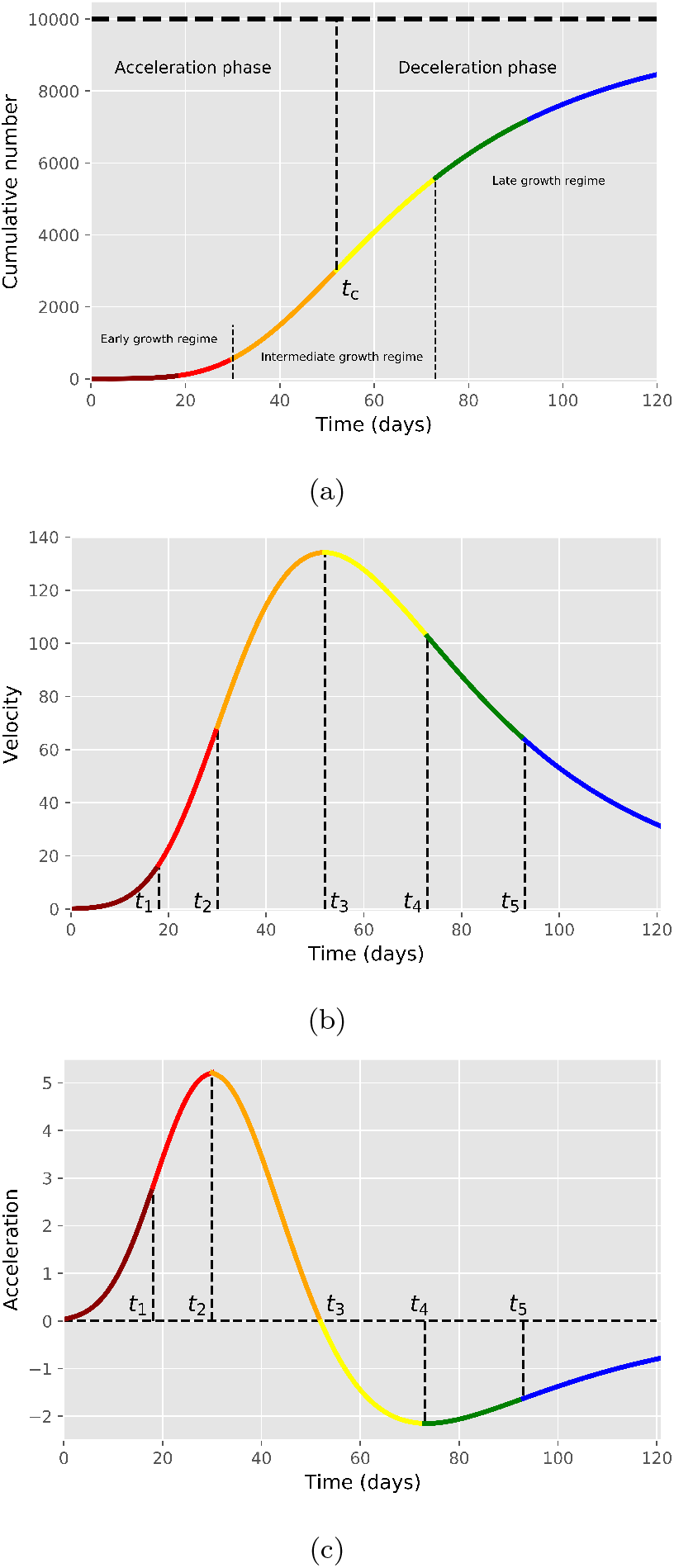
(a) Schematic of an epidemic curve for the cumulative number of deaths, with an indication of its two main phases, namely, the acceleration and deceleration phases, respectively. The point *t*_*c*_ indicates the inflection point of the cumulative curve, where the acceleration is zero, which separates the two phases. (b) Velocity and (c) acceleration curves, corresponding to the first and second derivatives, respectively, of the cumulative curve shown in (a). The different colors in the three curves indicate the respective epidemic stages, according to the corresponding acceleration regimes; see text.

These two phases are separated by the inflection point, *t*_*c*_, of the cumulative curve, which corresponds to the point where the acceleration is zero. In Fig. 1(b) we show the ‘velocity’ curve, *Ċ*(*t*), where dot denotes time derivative, which corresponds to the daily number of deaths. As illustrated in this figure, the inflection point, *t*_*c*_, of the cumulative curve corresponds to the “peak” of the daily curve, which separates the phases of increasing and decreasing growth velocity, respectively.

From a more practical viewpoint, a finer classification scheme of the different growth regimes can be given as follows: i) the *early growth regime*, where the acceleration is always increasing; ii) the *intermediate growth regime*, which is characterized by a nearly-linear profile around the inflection point; and iii) the *late growth regime*, for which the deceleration (i.e., the absolute value of the negative acceleration) is always decreasing. Note that in the late growth regime, the curve bends away from the linear regime and thus starts its approach toward the final plateau, represented by the horizontal dashed line in Fig. 1(a). Moreover, each of these three growth regimes can be further divided into two stages, on the basis on the respective trends of the acceleration curve in each regime, as described below.

In Fig. 1(c), we show the acceleration curve corresponding to the cumulative curve of Fig. 1(a). As shown in Fig. 1(c), the acceleration curve has five important characteristic points, namely: i) the inflection point, *t*_1_, in the region of positive acceleration; ii) the point, *t*_2_, of maximum acceleration; iii) the point, *t*_3_, of zero acceleration, which corresponds to the inflection point of the cumulative curve, i.e., *t*_3_ = *t*_*c*_, where 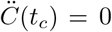; iv) the point, *t*_4_, of minimum acceleration, which corresponds to the point of maximum deceleration; and v) the inflection point, *t*_5_, in the region of negative acceleration. Recalling that the rate of change of the acceleration is known as the “jerk” (or “jolt”), we thus have that *t*_2_ and *t*_4_ are the points of zero jerk, meaning that 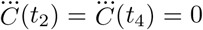; while *t*_1_ and *t*_5_ are the points of maximum jerk, i.e., 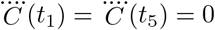.

In view of the existence of the five characteristic points, *t*_*i*_, *i* = 1, …, 5, discussed above, we can accordingly define six dynamical stages, as follows:

1. Increasing acceleration and increasing jerk (dark red stage): *t < t*_1_.
2. Increasing acceleration and decreasing jerk (light red stage): *t*_1_ *< t < t*_2_.
3. Decreasing acceleration (orange stage): *t*_2_ *< t < t*_3_.
4. Increasing deceleration (yellow stage): *t*_3_ *< t < t*_4_.
5. Decreasing deceleration and increasing jerk (green stage): *t*_4_ *< t < t*_5_.
6. Decreasing deceleration and decreasing jerk (blue stage): *t > t*_5_.

These six dynamical stages are indicated by the respective colors in the three curves shown in Fig. 1.

Note that the early growth regime has two dynamical stages, indicated by dark and light red in Fig. 1(a), respectively. In this regime, the acceleration is always increasing, see Fig. 1(c), with the two stages being differentiated only as to whether the jerk is increasing (dark red) or decreasing (light red).

From a practical viewpoint, however, the distinction between these two early growth stages is not very relevant, since the first stage is relatively short. Therefore, these two initial dynamical stages will be treated in our classification scheme (see below) as only one epidemic stage, namely that of increasing acceleration.

The intermediate growth regime comprises two epidemic stages, namely, the stage of decreasing acceleration (orange), followed by the stage of increasing deceleration (yellow), as seen in Fig. 1. Note that during the intermediate regime the growth profile is approximately linear, since the two corresponding stages are close to the inflection point, *t*_*c*_, at which the acceleration is zero, thus leading to a nearly linear growth around it.

Finally, the late growth regime corresponds to the regime of decreasing deceleration, which entails two dynamical stages, depending on whether the jerk is increasing (green) or decreasing (blue). As this regime begins, the cumulative curve starts to bend more strongly, thus moving away from the quasi-linear behavior of the intermediate regime. In this sense, the first stage (green) of the late growth regime marks the transition toward the saturation of the epidemic curve, and so we shall refer to this stage as transition to saturation. Similarly, the last stage (blue) corresponds to the saturation itself, where the first three derivatives of *C*(*t*) are all decreasing, and so it may be said that the epidemic is relatively under control once it enters this final stage. (If, however, control measures are relaxed prematurely, there may be a recrudescence of infections and the death curve may accelerate again, causing the so-called second wave of infections. These effects, however, will not be considered in this analysis).

Given a real epidemic curve at a given current time *t*_*f*_, it is important to determine in which dynamical stage the epidemic is at the chosen location at that time. To do so, it is first necessary to have a mathematical model that provides a theoretical curve describing the data, from which one can calculate the characteristic points *t*_*i*_, *i* = 1, …, 5, discussed above. Once those points have been obtained, one can then compare the position of the time *t*_*f*_ with respect to the characteristic points *t*_*i*_ obtained from the mathematical model. The stage of the epidemic can thus be determined according to the following classification scheme (and respective color code):

1. Stage of increasing acceleration (red): *t*_*f*_ *< t*_2_.
2. Stage of decreasing acceleration (orange): *t*_2_ *< t*_*f*_ *< t*_3_.
3. Stage of increasing deceleration (yellow): *t*_3_ *< t*_*f*_ *< t*_4_.
4. Transition to saturation (green): *t*_4_ *< t*_*f*_ *< t*_5_.
5. Saturation stage (blue): *t*_*f*_ *> t*_5_.

The `ModInterv` system, to be discussed below, implements four mathematical growth models, with different levels of complexity, from which the dynamical stage of any given epidemic curves (with no second-wave effects) can be determined.

### 3.2 Mathematical Models

The most general model, of which the others are particular cases, is the beta logistic model (BLM), described by the following ordinary differential equation [8, 11]:

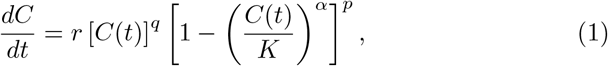

where *r* is the growth rate in the initial stage, *K* is the final size of the epidemic, *q* controls the initial growth regime and allows interpolar from linear growth (*q* = 0) to subexponential growth (*q <* 1) to purely exponential growth (*q* = 1), The exponent *p* controls the approach rate to the plateau, with *p >* 1 implying a polynomial rate, while *p* = 1 produces an exponentially fast approach, and exponent *α* is the so-called asymmetry parameter. BLM has as particular cases of interest the following models: the generalized Richards model (GRM), when *p* = 1; the Richards model (RM), for *q* = *p* = 1; and the q-exponential model, in case *p* = 0. More details about the above models can be found in the references [8, 11, 12].

BLM also retrieves the standard logistic model, in which case *q* = *p* = *α* = 1, and therefore Eq. (1) is also known as the generalized logistic equation [8]. It is worth remembering, however, that the logistic model describes a symmetric growth curve in relation to the inflection point. But since this symmetry is rarely observed in the empirical curves of COVID-19, this particular case is not implemented separately in `ModInterv`. Note, however, that in the case of the Richards model, where *q* = *p* = 1, the exponent *α* is free to vary, so that a symmetric curve can in principle be obtained if the numeric adjustment returns the value *α* = 1.

The BLM admits an analytic solution [11] in the following implicit form:

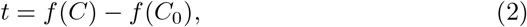

where

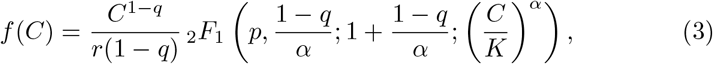

with _2_*F*_1_(*a, b*; *c*; *x*) being the Gauss hypergeometric function.

The inflection point of the BLM curve, *C*(*t*), is located at the time *t*_3_ = *f* (*Kx*_3_), where

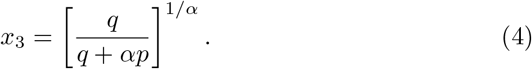

Two other important characteristic points of the curve *C*(*t*) are the points of maximum and minimum acceleration, denoted by *t*_2_ and *t*_4_, respectively; see Fig. 1(b). Note that since the 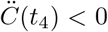, the point *t*_4_ corresponds in fact to the point of maximum deceleration (i.e., maximum value of 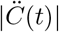 within the region of negative acceleration). Recalling that the rate of acceleration is known as the jerk (or jolt), we thus have that the points *t*_2_ and *t*_4_ are the points of zero jerk, which can be obtained from the condition 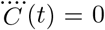. After some tedious algebra one finds that *t*_2,4_ = *f* (*Kx*_2,4_), where

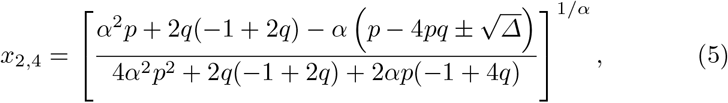

with Δ being given by

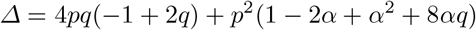

One can also compute the points *t*_1_ and *t*_5_ of maximum jerk, i.e., 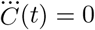. But in this case the expressions are prohibitively long and will not be given here.

Setting *p* = 1 in Eqs. (3)–(5) yields the exact solution for the GRM, together with the corresponding expressions for its characteristic points *t*_2_, *t*_3_, and *t*_4_. One important distinction between the general BLM (i.e., with *p >* 1) and the GRM (*p* = 1) is that the former predicts a polynomial approach to the plateau [11]:

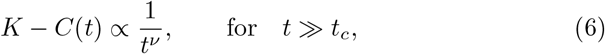

where *ν* = 1*/*(*p* −1). In contradistinction, the GRM describes an exponentially fast approach to the plateau:

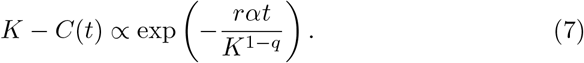

Furthermore, for *p* = *q* = 1, the BLM reduces to the RM, as already mentioned, in which case one obtains an explicit solution in the following form [12]:

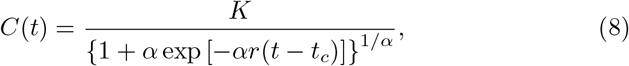

where the inflection point *t*_*c*_ can be obtained in terms of the initial condition *C*_0_ via the relation: *C*_0_ = *K/*[1 + *α* exp (*αrt*_*c*_)]^1*/α*^. For the RM, the points, *t*_2_ and *t*_4_, of maximum acceleration and maximum deceleration, can be obtained explicitly by solving 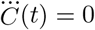, which yields

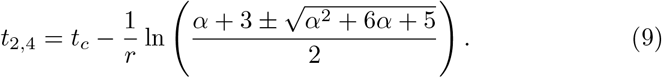

Again, the expressions for the points *t*_1_ and *t*_5_ of maximum jerk are rather long and will not be given here.

When the outbreak in a given place is in its early stage, i.e., when the epidemic curve is still in a regime of increasing acceleration, see Fig. 1(b), none of the previous models (RM, GRM and BLM) are suitable, because the parameters estimated from these three models (if they converge at all) are not reliable. More specifically, as the empirical curve does not exhibit yet an inflection point, estimating the parameter *α* becomes unreliable. In this case, it is preferable to use the so-called generalized growth model [2, 12], which is defined by setting *C ≪ K* in (1), which results in the following ODE:

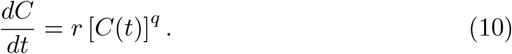

The solution of (10) is

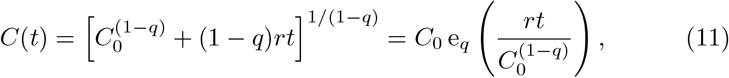

where the function e_*q*_(*x*) = [1 + (1 −*q*)*x*]^1*/*(1−*q*)^ is known in the physics literature as the *q*-exponential function [7]. For this reason we shall also refer to model (10) as the *q-exponential model*. In particular, note that according to (11) the growth of the epidemic curve in this regime is approximately polynomial (for 0 ≤ *q <* 1) as *t* increases:

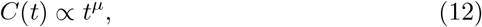

where *µ* = 1*/*(1− *q*); whereas only for *q* = 1 does one obtain an exponential growth. Polynomial early growth has been identified in the COVID-19 data for several countries [14, 5, 12].

The application `ModInterv` implements the four growth models discussed above, namely, BLM, GRM, RM, and *q*-exponential. More specifically, for a given epidemic curve, the system initially tries to adjust the empirical data with the BLM. In general, however, the convergence of this model only happens for curves that already are in a saturation phase, which allows estimating the exponent *p* that controls the approach to the plateau. If there is no convergence of BLM, the `ModInterv` next tests the GRM. As already mentioned, the GRM assumes an exponential behavior in the saturation phase, but still requires the numerical adjustment of two exponents: the exponent *q*, which controls the subexponential growth at the beginning, and the exponent of asymmetry *α*. For this reason, the GRM only produces a good adjustment when the curve is already in the deceleration phase. In cases where the GRM does not converge satisfactorily, then the we apply the RM. Since the RM assumes exponential behavior both at the beginning and at the end of the curve, this model requires the adjustment of only the asymmetry exponent *α* (in addition to the parameters *r* and *K*), being therefore the most robust model regarding convergence. In general, the RM is more suitable for epidemic curves in an intermediate stage, but it may also apply to curves in more advanced stages for which neither the BLM nor the GRM are suitable.

Once the theoretical model (among the BLM, GRM and RM) that best describes the data is determined, the `ModInterv` computes the characteristic points *t*_*i*_ and thus determines the stage of the epidemic in that locality, according to the classification scheme described above.

If, however, the `ModInterv` identifies an empirical curve in the stage of increasing acceleration, the fitting of the data is redone with the *q*-exponential model given in (11). This is necessary because during the early growth regime the empirical curve is still far from developing an inflection point, so that the theoretical prediction for *t*_*c*_ obtained from the RM or the GRM will lie well beyond the last date *t*_*f*_ and hence this estimate for *t*_*c*_ is not reliable. More to the point, in such case there are not enough data yet to yield a reliable estimate of the parameter *α*, and hence it is preferable to use the *q*-exponential model, which describes only the initial growth.

Once the model that best describes a given empirical curve has been identified, the `ModInterv` app displays the figures of the cumulative and daily curves, showing the empirical data together with the respective theoretical fits. The figure title identifies the location in question and informs the model that best adjusted the curve, while the legend box gives the fitting parameters as well as the epidemic stage of that location.

### 3.3 Numerical Analysis

All numerical data fittings implemented in the `ModInterv` are performed via Levenberg-Marquardt (LM) algorithm to solve the corresponding non-linear optimization problem by the least squares method. The computational codes to perform the respective fits are written in the `Python` language and employ the LM algorithm implemented in the lmfit package for `Python`, which has a built-in routine for estimating the errors of the fitted parameters via the covariance matrix [6].

In the `ModInterv`, the results of the fitting procedure are deemed acceptable when the errors in the parameters are smaller than a certain threshold. For the analysis presented here the error tolerance was set at 20% of the values estimated for the parameters themselves.

## 4 RESULTS

In Fig. 2, we show the map of Brazil, in which each state and the Federal District are painted with the color corresponding to the respective stages of the COVID-19 epidemic on August 21, 2020. As already mentioned, this date was chosen because it corresponds to a period in which all five stages of the COVID19 epidemic are represented in Brazil. The maps shows that distribution of the epidemic in a continental country like Brazil is very heterogeneous. Below we discuss this situation in more detail.

**Fig. 2:**
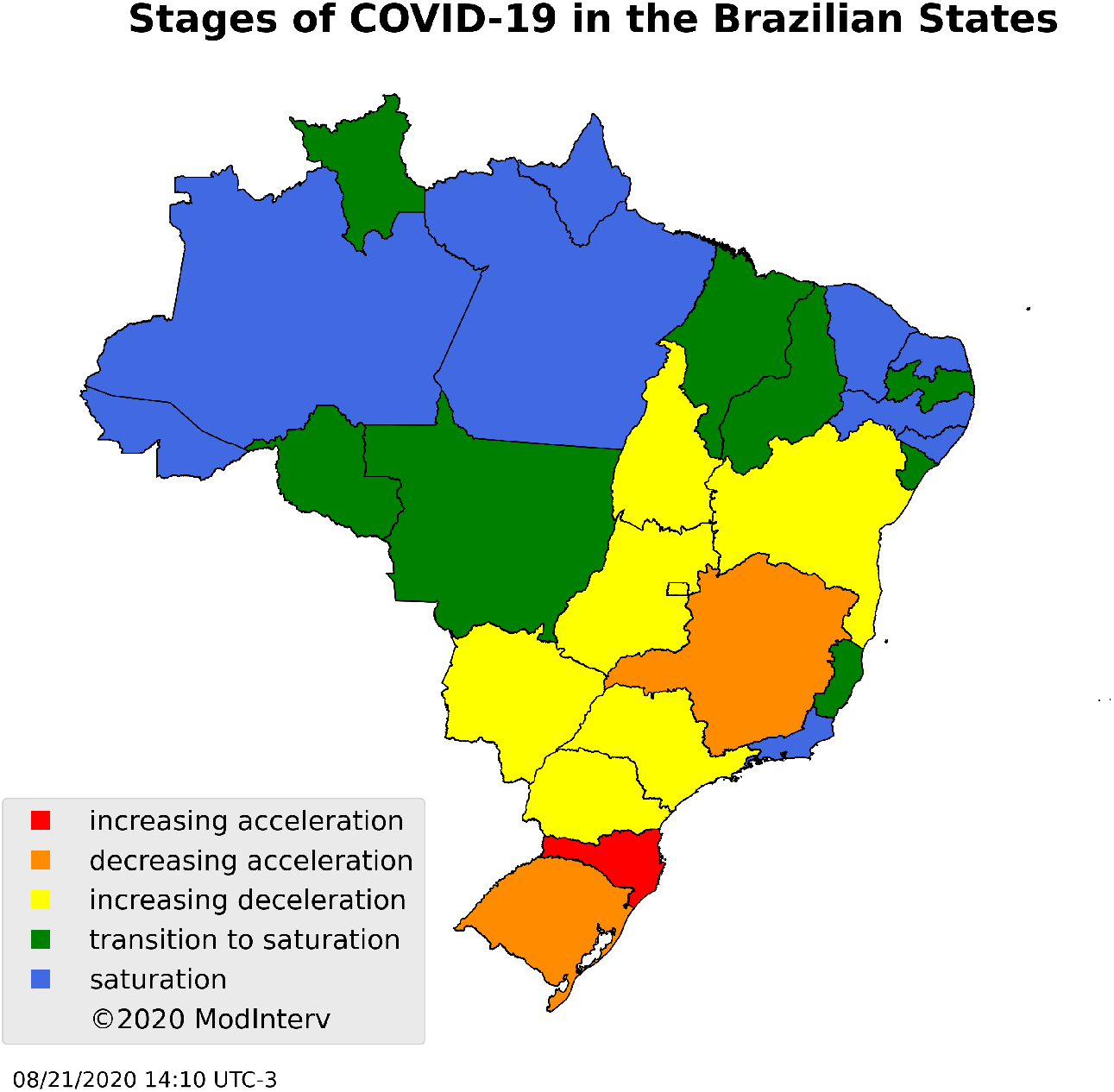
Situation of the COVID-19 epidemic in the Brazilian States and Federal District, on August 21, 2020, according to the classification of the epidemic stages implemented in the `ModInterv` application.

In Figs. 3–10, we present the plots produced by the `ModInterv` for each of the Brazilian federal units, organized into groups according to the respective stages of the epidemic. In the left panel of each figure we present the empirical data (red circles) and the theoretical curves (black curve) for the cumulative number of deaths attributed to COVID-19 for the respective federal unit.

**Fig. 3:**
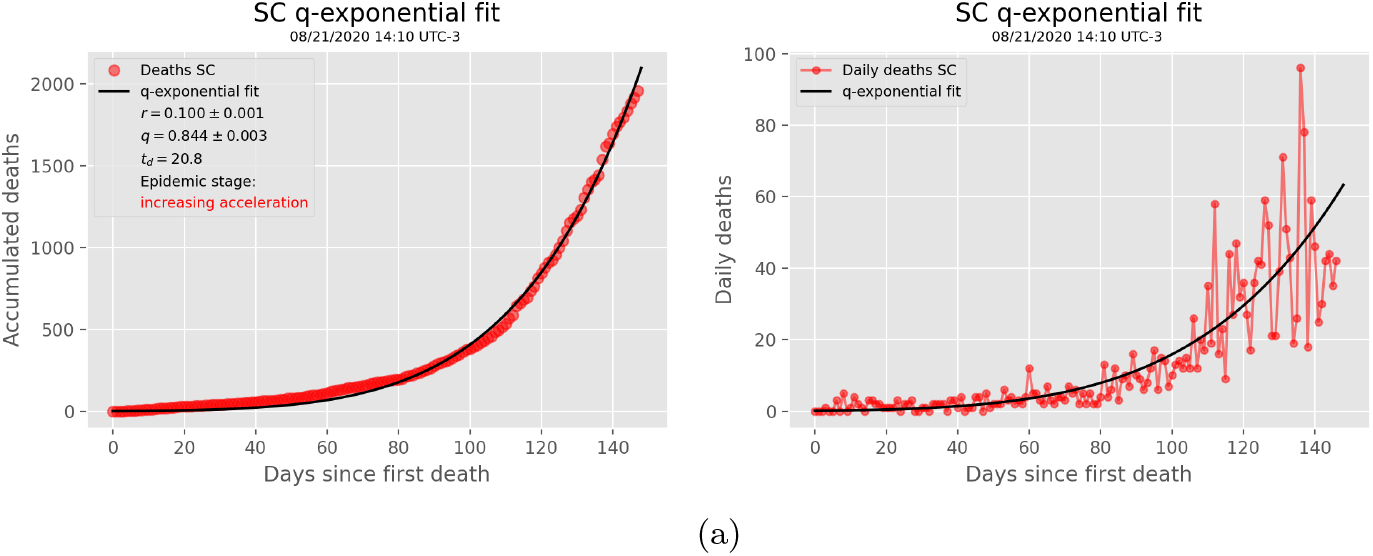
State in the stage of increasing acceleration.

In Figs. 3 the right panel show the corresponding daily number of deaths, together with the derivative *dC/dt* of the theoretical curve *C*(*t*) shown in the respective left panel. Below we discuss in more details the results for each of the respective groups.

### 4.1 State in the stage of increasing acceleration

The empirical curve for the state of Santa Catarina is best adjusted by the *q*-exponential growth model, as shown in the left panel of Fig. 3, indicating that in this state the COVID-19 fatality curve due was still in a dynamical regime of increasing acceleration. In the right panel of Fig. 3, we show the daily number of deaths, together with the derivative of the theoretical curve, where we can see that the agreement between data and theory is also very good for the daily data.

From Fig. 3, we observe a linear growth, which goes approximately up to 60th day after the first death. This shows that in the first two months the epidemic curve for Santa Catarina exhibited a regime of low growth (i.e., with near zero acceleration, i.e. *q* ≈ 0), as a reflection of the non-pharmacological intervention measures adopted at the beginning of the pandemic. Later, with the premature flexibilization of these measures, the curve quickly changed trajectory, passing to exhibit an almost exponential growth (i.e., with *q* close to 1).

### 4.2 States in the stage of decreasing acceleration

The states of Minas Gerais and Rio Grande do Sul are in the first stage of the intermediate growth regime, namely the stage of decreasing acceleration, as shown in Fig. 4. It can be seen from this figure that the last point of each of the empirical curves had already exceeded the orange line, corresponding to the point *t*_2_ of maximum acceleration, but there had not yet appeared the formation of the inflection point in the empirical curve, thus characterizing the second epidemic stage, as defined before.

**Fig. 4:**
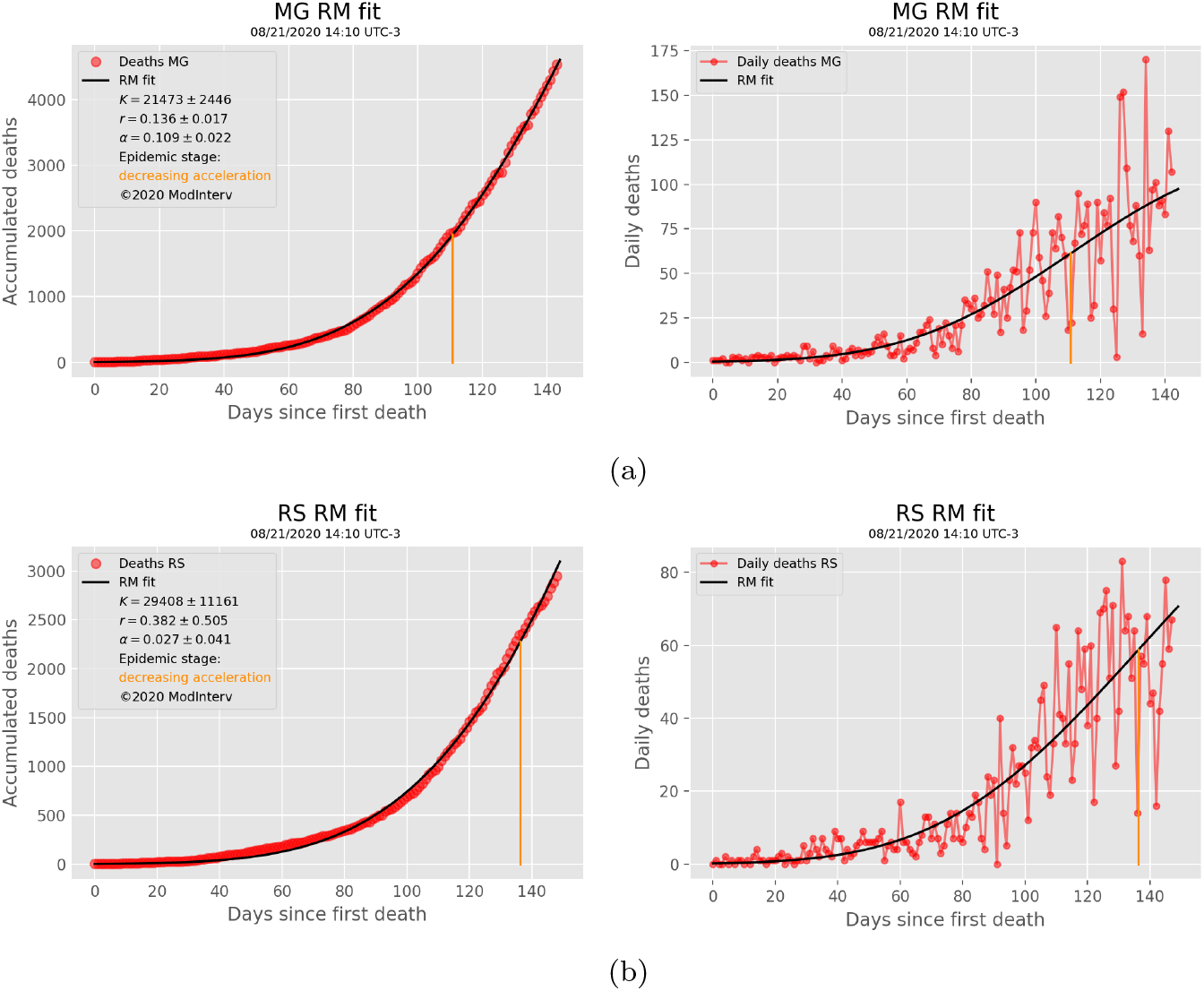
States in the stage of decreasing acceleration.

One can also see from Fig. 4 that the epidemic curves for the states of Minas Gerais and Rio Grande do Sul exhibit a sudden change in trend around the second month of the epidemic, passing from a linear regime to a quasi-exponential growth. This quick change in trend is a similar to what happened to the epidemic curve of Santa Catarina around the same time (see discussion above) and is probably due to the same reasons (i.e., premature relaxation of the control measures). However, at the moment of the present analysis, the states of Minas Gerais and Rio Grande do Sul had already left the regime of increasing acceleration (where Santa Catarina still was) and progressed to the stage of decreasing acceleration.

### 4.3 States in the stage of increasing deceleration

As seen in Figs. 5–6, the states of Bahia, Goiás, Mato do Grosso do Sul, Paraná, São Paulo, Tocantins, and Distrito Federal are in the second stage of the intermediate phase, which is characterized by an increasing deceleration. The mathematical fits suggest that the epidemic curves for these states had already passed through the inflection point (indicated by the yellow line), and so they had started the deceleration phase of the epidemic, but they had not yet reached the point *t*_4_ of maximum deceleration, hence the epidemic curves had not entered the late growth regime.

**Fig. 5:**
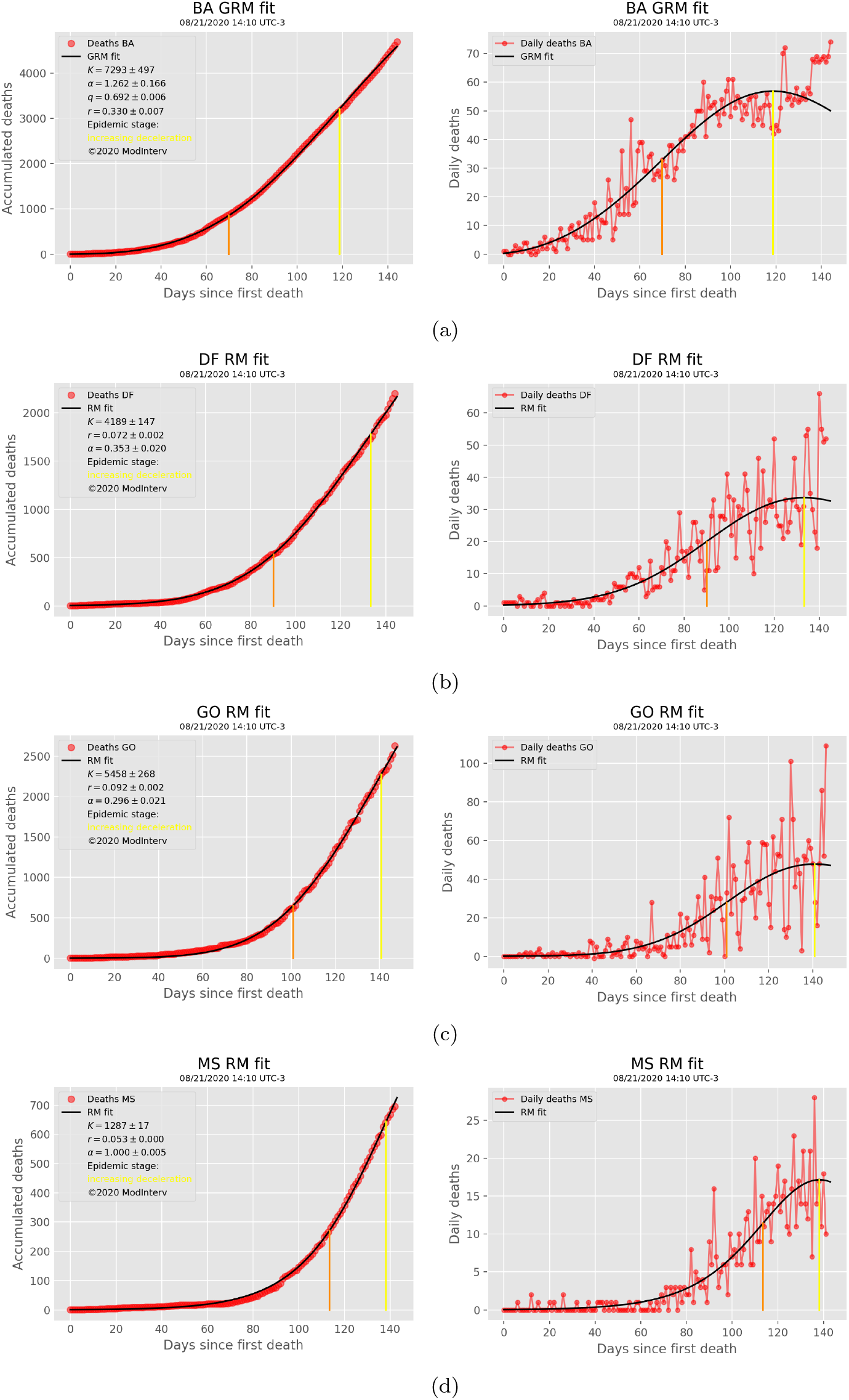
States in the stage of increasing deceleration - I.

**Fig. 6:**
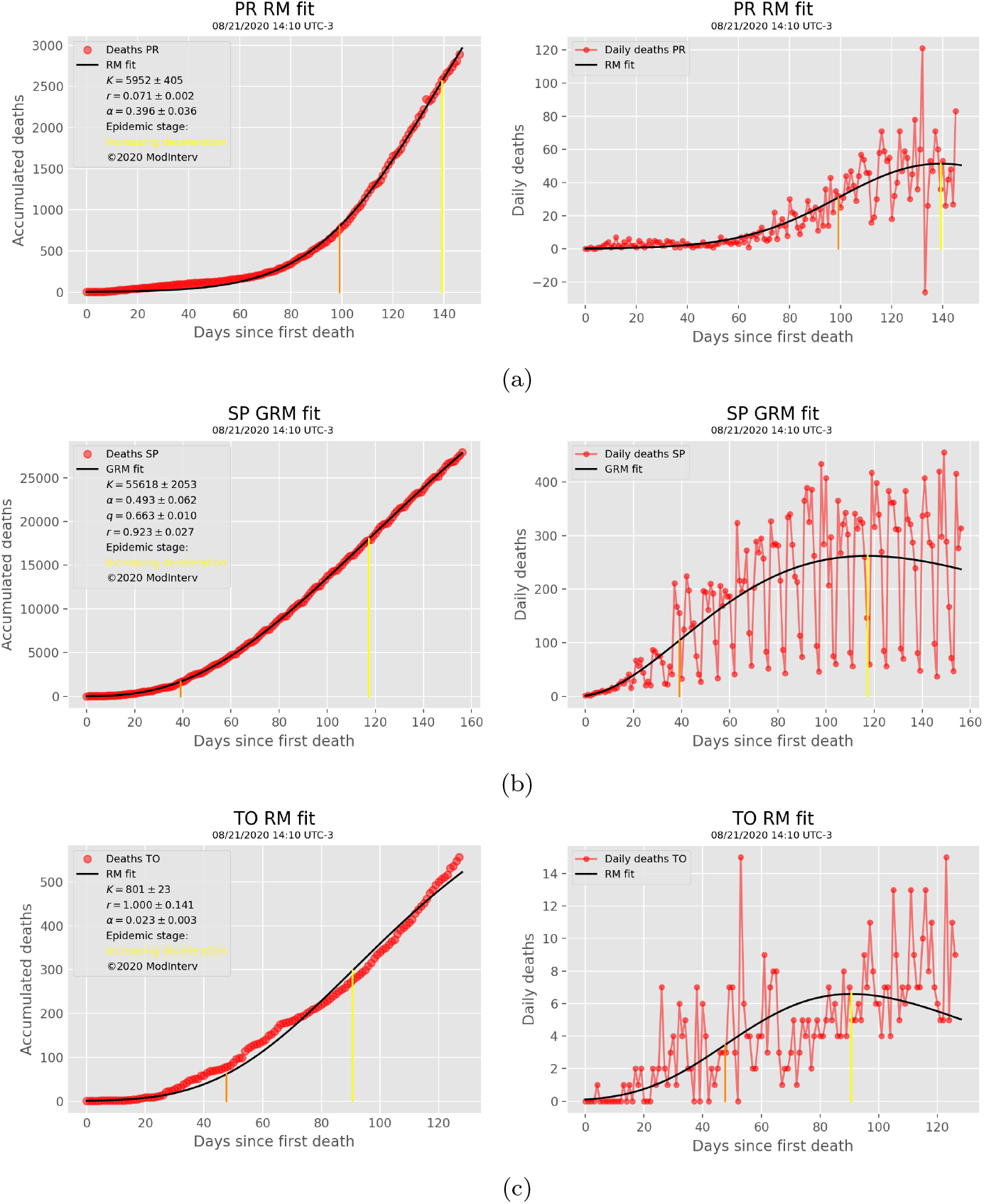
States in the stage of increasing deceleration - II.

### 4.4 States in the stage of transition to saturation

The states of Espírito Santo, Maranhão, Mato Grosso, Paraíba, Piauí, Rondônia, Roraima, and Sergipe were found to be in the stage of transition to saturation, as shown in Figs. 7–8. As previously discussed, this stage is the first stage of the late growth regime and is characterized by the fact that the epidemic curve has already passed the point *t*_4_ of maximum deceleration (green line).

**Fig. 7:**
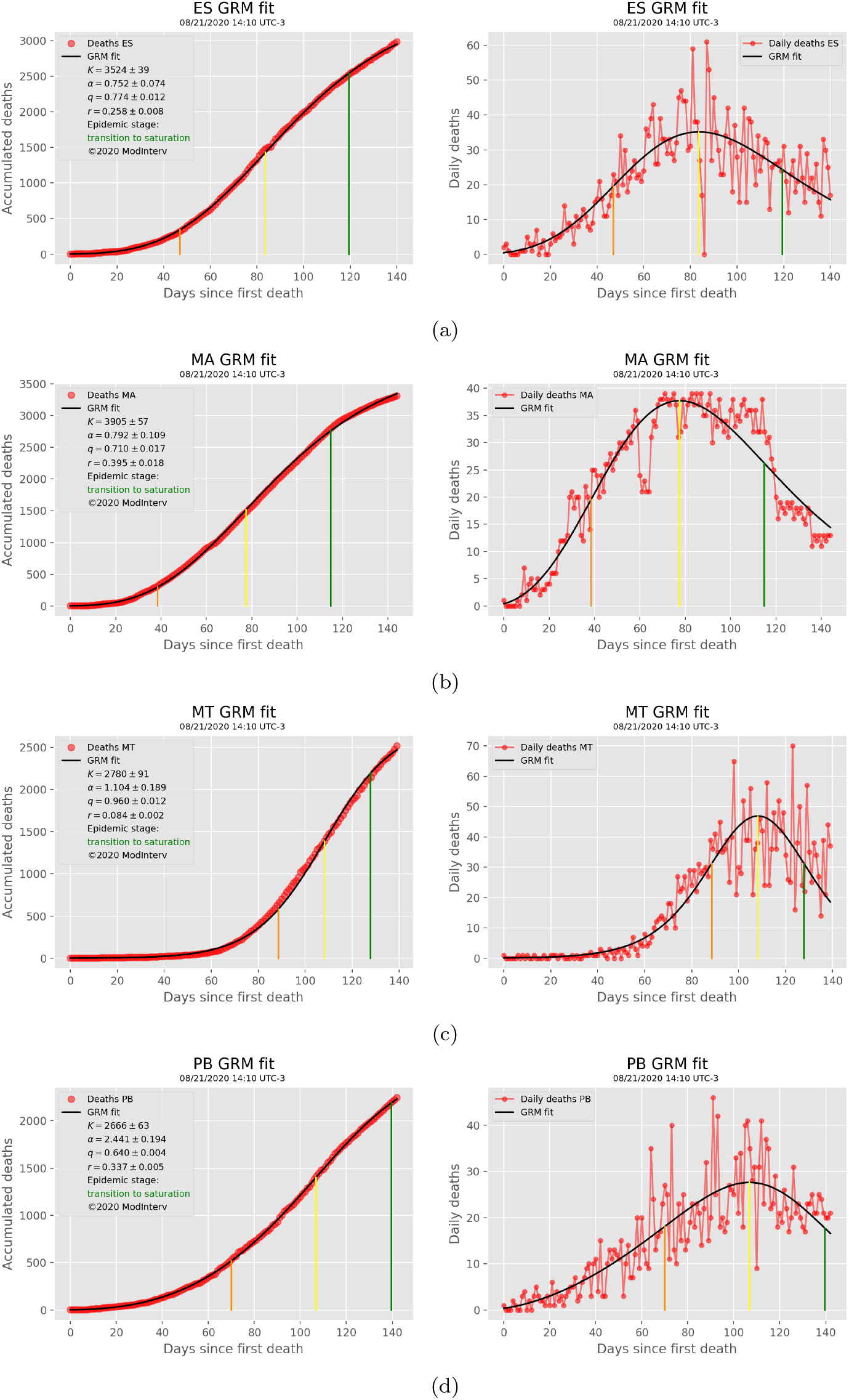
States in the transition to saturation stage - I.

**Fig. 8:**
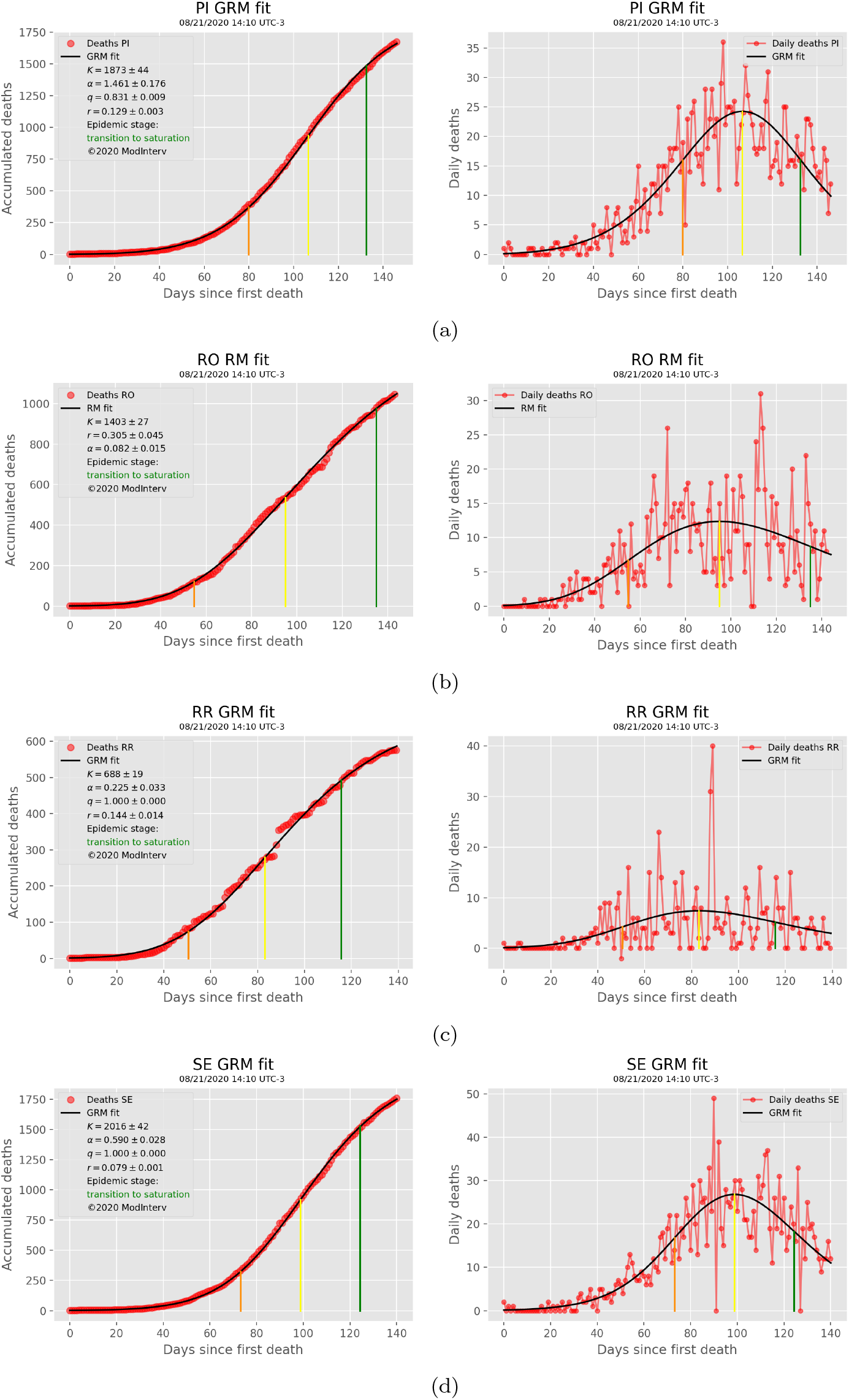
States in the transition to saturation stage - II.

Therefore, so it is already in a regime of decreasing deceleration but still with an increasing jerk. This growing jerk forces the curve to deviate away from the near-linear growth of the intermediate phase, thus contributing to the initial bending of the epidemic curve. We can thus say, with a certain degree of reliability, that for the states at this stage that the epidemic (or rather its first wave) was beginning to get under control.

### 4.5 States in the saturation stage

Finally, we have the states of Acre, Alagoas, Amazonas, Amapá, Ceará, Pará, Pernambuco, Rio de Janeiro, and Rio Grande do Norte, which are already in the stage of saturation proper, as can be seen in Fig. 9–10. At this stage, the epidemic curve has already surpassed the blue line, which corresponds to the point *t*_5_ of maximum jerk. Therefore, at this last stage of the epidemic, the velocity, acceleration, and jerk of the epidemic curve are all decreasing functions of time, so that the epidemic is approaching its final plateau. Therefore, it may be said that at this stage the epidemic is relatively under control (assuming, of course, that the saturation trend is maintained over time).

**Fig. 9:**
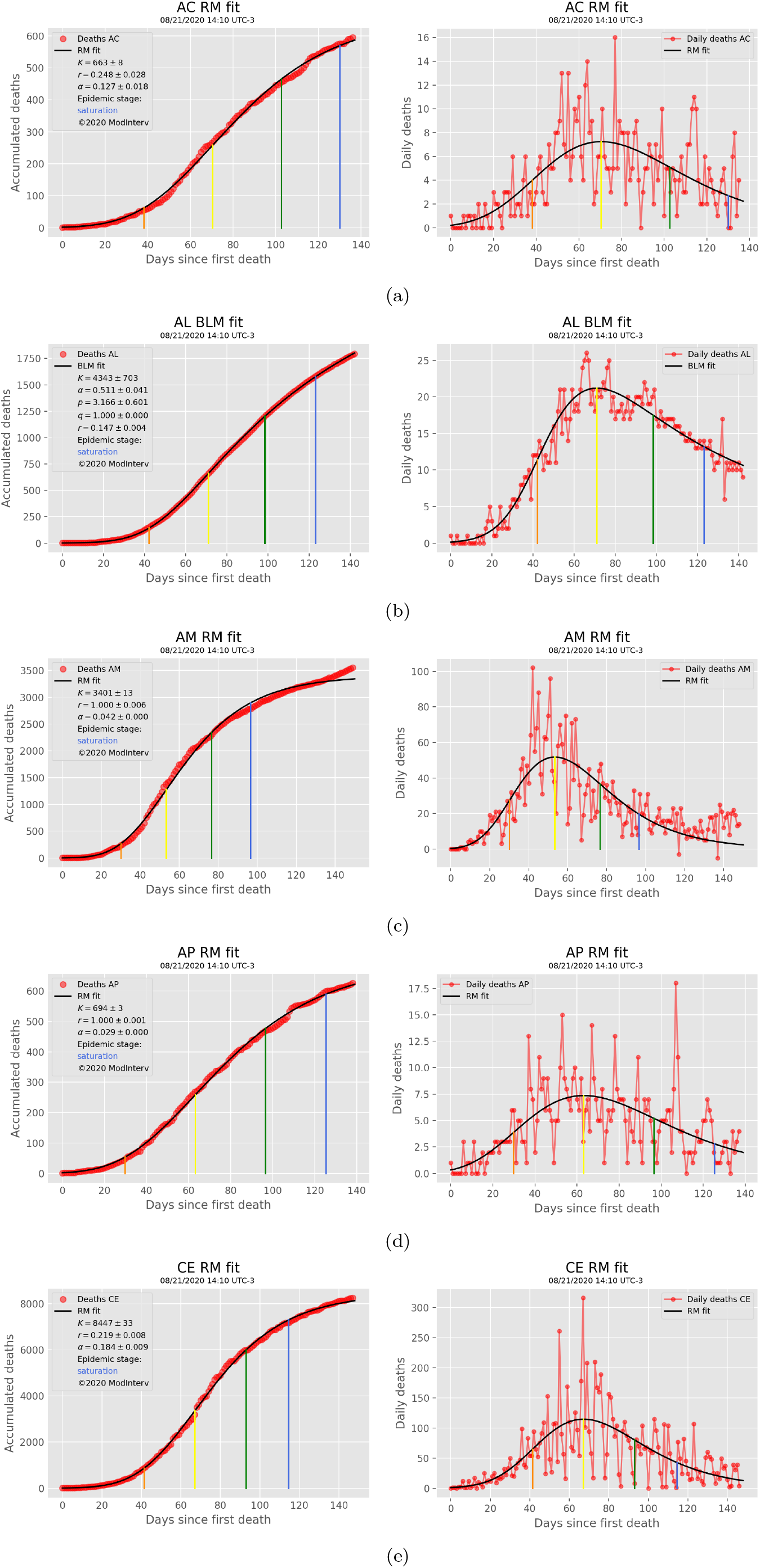
States in the saturation stage - I

**Fig. 10:**
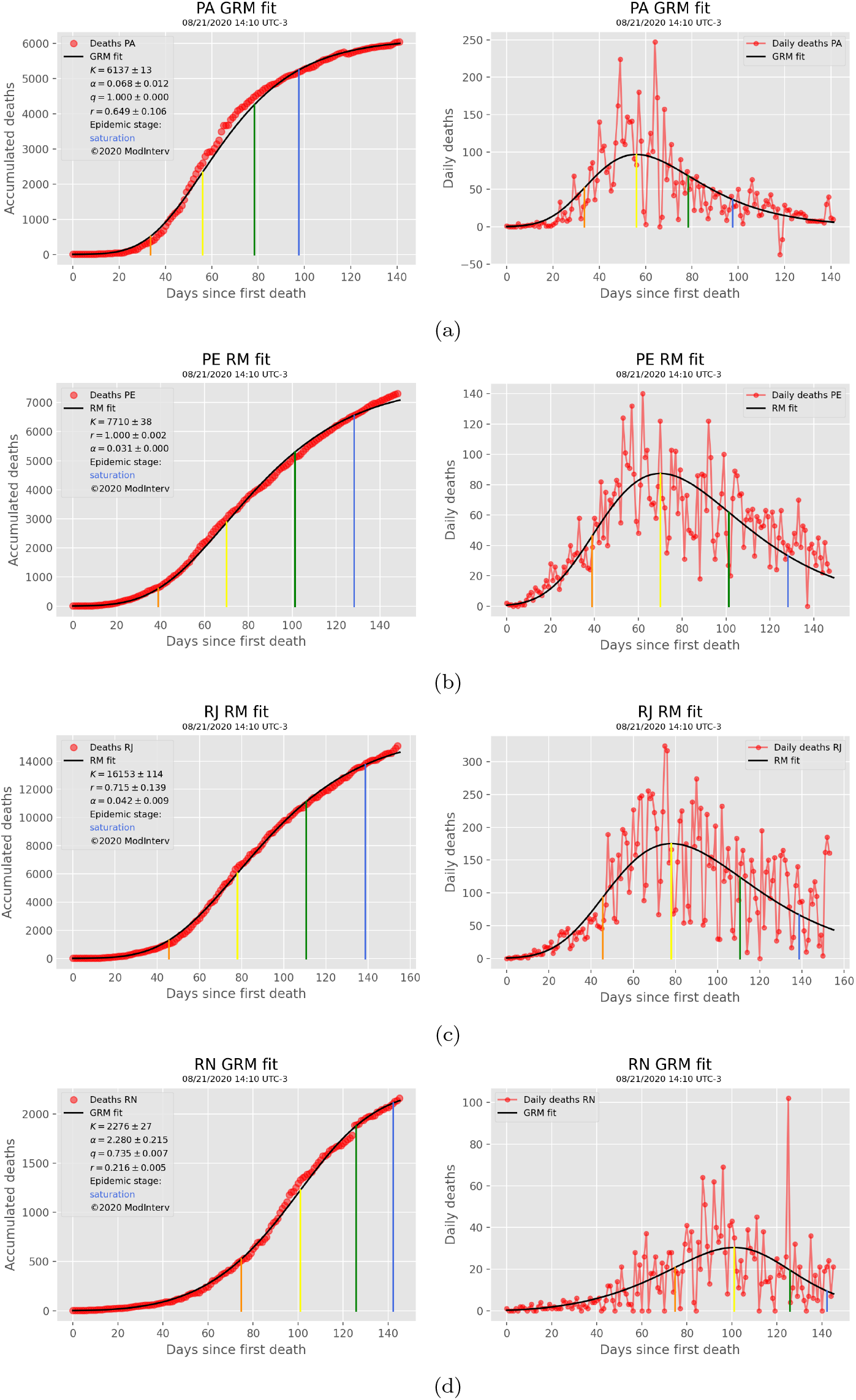
States in the saturation stage - II

## 5 Discussion

In this work we have applied four mathematical growth models, implemented in the `ModInterv` application, to study the COVID-19 fatality curves of the Brazilian States and the Federal District, up until August 21, 2020. Up to that date, nearly five months after the first death in Brazil, all five epidemic stages were represented in at least one state throughout the country.

We verified, in particular, that Santa Catarina was the only state in the early growth regime, where the epidemic curve was still exhibiting an increasing acceleration. The other two states of the Southern Region, Rio Grande do Sul and Paraná, had recently emerged from this early growth regime and were already in the first and second stages of the intermediate growth regime, corresponding to the stages of decreasing acceleration and increasing deceleration, respectively. Thus, all three Southern states were at that time far from curtailing the epidemic.

In contradistinction, all states in the Northern and Northeastern Regions, except for Bahia and Tocantins, were either in stage of transition to saturation (MA, PB, PI, RO, RR, and SE) or in the saturation stage (AL, AM, AP, CE, PE, and RN), which are the two final stages of the epidemic (if there is no recrudescence of infections). Bahia and Tocantins were in the stage of increasing deceleration, but had not yet reached the transition to the saturation stage.

Among the Midwestern states, only Mato Grosso had already started the transition to saturation, while Goiás, Mato Grosso do Sul and the Federal District were still in the stage of increasing deceleration. The Southeastern region had the highest diversity of epidemic stages, in the sense that each state was in a different stage, namely: Minas Gerais was in the stage of decreasing acceleration, São Paulo was in the stage of increasing deceleration, Espírito Santo was in the stage of transition to saturation, and Rio de Janeiro was the only Southeastern state which was already in the saturation stage.

The present analysis allows the interesting conclusion that five months onto the COVID-19 epidemic in Brazil the Northern and Northeastern regions were, in general, in a relatively more advanced phase of control of the pandemic; whereas in the more Southern states the epidemic was still accelerating or displayed only a slight deceleration. This general overview of the COVID-19 situation in Brazil largely reflects the distinct evolution patterns of the epidemic in the different regions of the country. For example, in many capitals of the Northern and Northeastern states, there was a strong accelerated growth in the number of deaths in the first few months of the epidemic, which forced local authorities to adopt more drastic measures to contain the spread of the virus. These measures contributed to the subsequent “bending” of the epidemic curve that we have seen in our analysis for these states. In contrast, in the Southern states, the evolution of the disease happened somewhat in the reverse: the initial growth of the epidemic curve was slow because of the early mitigation measures; but later on these measures were prematurely relaxed, which led, in turn, to a resurgence of the epidemic. As a consequence of this “relapse” of the epidemic, most Southern states were still in a phase of acceleration or, at most, of mild deceleration at the time of the present analysis.

Other states that presented this “relapse effect” were Goiás, Mato Grosso do Sul and Minas Gerais, meaning that these states also had a slow early growth in the first couple of months (nearly with a constant acceleration, i.e., *q* ≈ 0) but then experienced a quick surge in the number of COVID-19 deaths. As a result, by mid-August, 2020, they were far from the saturation phase, with Minas Gerais still in the decreasing acceleration phase and Goiás and Mato Grosso do Sul in the increasing deceleration phase. Another interesting example is the state of São Paulo, which maintained mitigation measures for a considerable period of time. These measures caused the curve to leave the stage of increasing acceleration relatively early, see Fig. 6(b), but were not sufficient to “bend” the curve and interrupt the almost linear of the intermediate growth regime, so that the state still remained in the stage of increasing deceleration.

It should be emphasized that the mathematical analysis presented here revealed the evolution of the COVID-19 epidemic in Brazil up to the maximum date (August, 21, 2020) considered in our analysis. The trajectories of the epidemic in the Brazilian federal units after that date have been strongly influenced by the maintenance or relaxation of control measures in the respective states. An analysis of the “full history” of the COVID-19 epidemic in Brazil, up to the present time, is beyond the scope of this paper. But a few comments are in order. For example, by October 9, 2020, all states had progressed to the late growth regime, i.e., they were either in the transition to saturation or in the saturation stages, indicating that the “first wave” of the epidemic was by then getting under control in Brazil. However, in late October and early November, 2020, the epidemic curves of several states started to show an upsurge, thus deviating from the up-to-then saturation trend. Such a recrudescence of the epidemic was likely the consequence of the so-called “second wave” of infections [10]. These effects will be studied later, since they require the use of more complex models with time-dependent parameters.

## Data Availability

The data used in our analysis are publicly available from the website https://covid19br.wcota.me, which compiles and updates the data from the bulletins published by the Health Departments of the Brazilian States.

## Acknowledgments

An early version of this paper was presented at the XXIII Congresso Brasileiro de Automática (CBA 2020). This work was partially supported by the National Council for Scientific and Technological Development (CNPq) in Brazil, through the grants Nos. 303772/2017-4 (GLV), 312612/2019-2 (AMSM), and 305305/2019-0 (RO). AAB also thanks CNPq for its support through a PhD Fellowship (grant No. 167348/2018-3). RO thanks D.A.D.O. and CASTLab.

